# Face masks for preventing respiratory infections in the community: A systematic review

**DOI:** 10.1101/2020.12.16.20248316

**Authors:** Maija Saijonkari, Neill Booth, Jaana Isojärvi, Jenni Finnilä, Marjukka Mäkelä

## Abstract

**Background:** The Ministry of Social Affairs and Health in Finland commissioned this systematic literature review on the effectiveness and safety of using face masks in public environments in protecting against upper respiratory tract infections, to inform policy. Previous reviews have not clearly distinguished the context of mask use.

**Methods:** The review was completed within two weeks, adhering to the PRISMA guidelines where possible. The review looks at the effect of face coverings (surgical masks or cloth coverings, excluding FFP2 and FFP3 masks) in protecting against the transmission in droplet-mediated respiratory tract infections. Our review includes controlled trials or previous systematic reviews of mask use by the general public in public spaces, outside homes and healthcare facilities.

**Results:** The systematic literature search identified five randomized trials. Use of masks prevented infections in one subgroup of one pilot study, so the effect of face masks on the transmission of infections outside the home appears small or nonexistent. Five of the eight systematic reviews showed no evidence of face masks being helpful in controlling the spread of respiratory infection or preventing exposure in healthy individuals. Meta-analyses often combined very heterogeneous studies and costs were not reported in any studies.

**Conclusions:** Randomized studies on the effect of face coverings in the general population are few. The reported effect of masks used outside the home on transmission of droplet-mediated respiratory infections in the population is minimal or non-existent. It is difficult to distinguish the potential effect of masks from the effects of other protective measures.

**Summary box:** *What is already known on this subject?:* Previous reviews on the effectiveness and safety of use of face masks in protecting against upper respiratory tract infections have not clearly distinguished the context of mask use. They have combined very heterogeneous studies done in homes, health care settings, or public environments.

*What does this study add?:* Our systematic review on the use of face masks in public environments, done to inform an impending policy decision, found five randomized trials (RCTs) and eight reviews. Use of masks prevented infections in one subgroup of one RCT, so the effect of face masks appears small or nonexistent.

## Background

The COVID-19 epidemic started in Finland in March 2020 and the government responded promptly by closing schools, promoting hand hygiene and social distancing. At the time of writing the use of masks has not been recommended in Finland – a policy similar to all other Nordic countries. In May, the Ministry of Social Affairs and Health (MSAH) asked us to prepare a systematic literature review on the effectiveness and safety of use of face masks in public environments in protecting against upper respiratory tract infections, to inform an impending policy decision, as previous reviews were unclear on this issue.

## Methods

The PRISMA guideline (1) was applied, with the study question (PICO) defined as follows: “What is the effect of face coverings (surgical masks or cloth coverings, excluding FFP2 and FFP3 masks) used by the population in public spaces outside or inside (excluding health care facilities and homes) in protecting against the transmission in droplet-mediated respiratory tract infections, such as COVID-19 or influenza?”

### Literature search

The systematic literature search carried out on 8 May 2020 found 108 scientific publications to be evaluated (Figure 1). We conducted a rapid, systematic search of MEDLINE (OvidSP), Cochrane Database of Systematic Reviews and Cochrane Central Register of Controlled Trials; our search strategies are presented in Appendix 1. We also searched the reference lists of the studies and reviews included. Information of which randomized studies meeting the inclusion criteria for this review were included in previous reviews illustrates overlaps (Table 1).

**Table 1.**
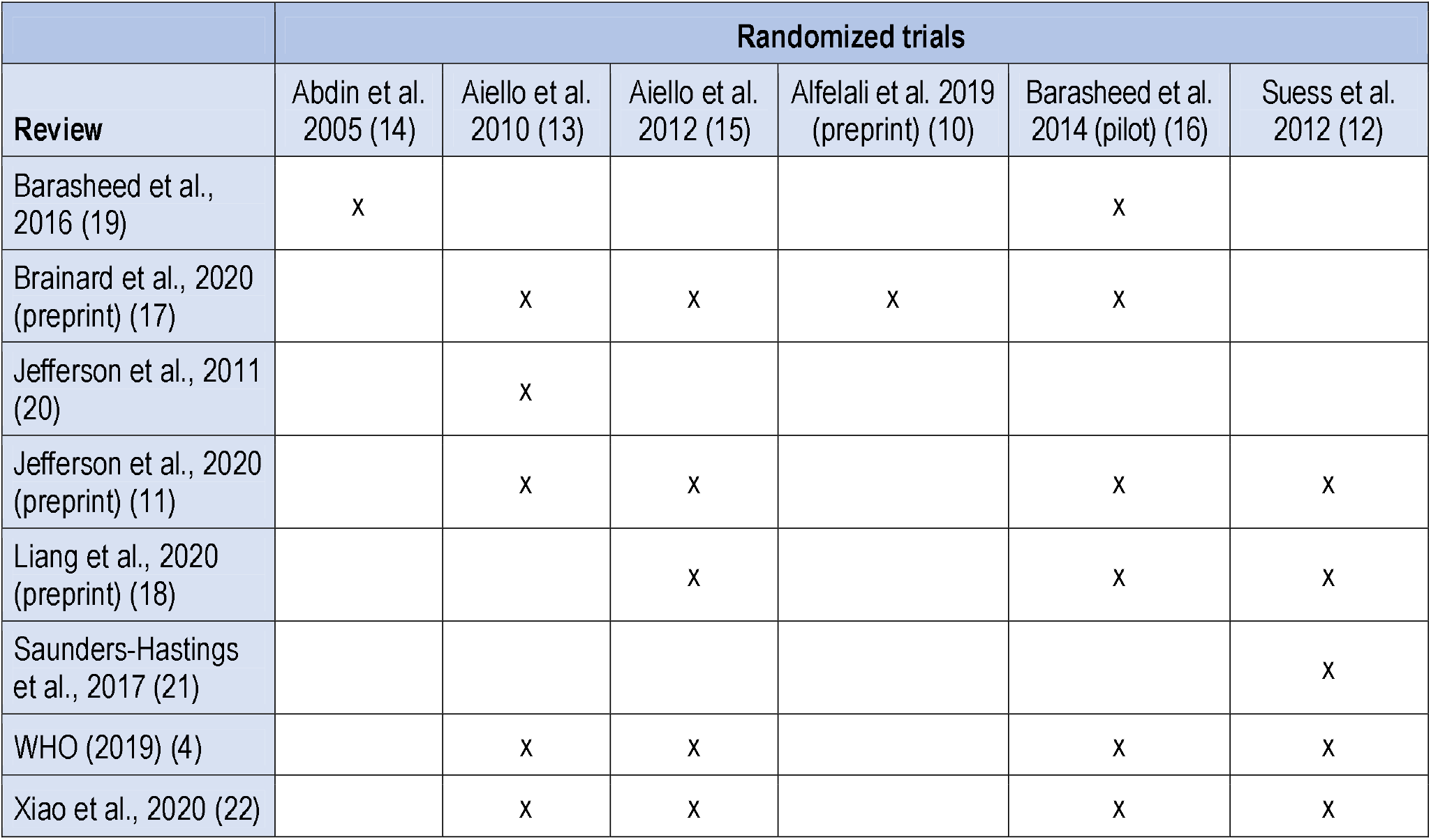
Randomized trials on face mask use in public environments included in previous systematic reviews.

**Figure 1:**
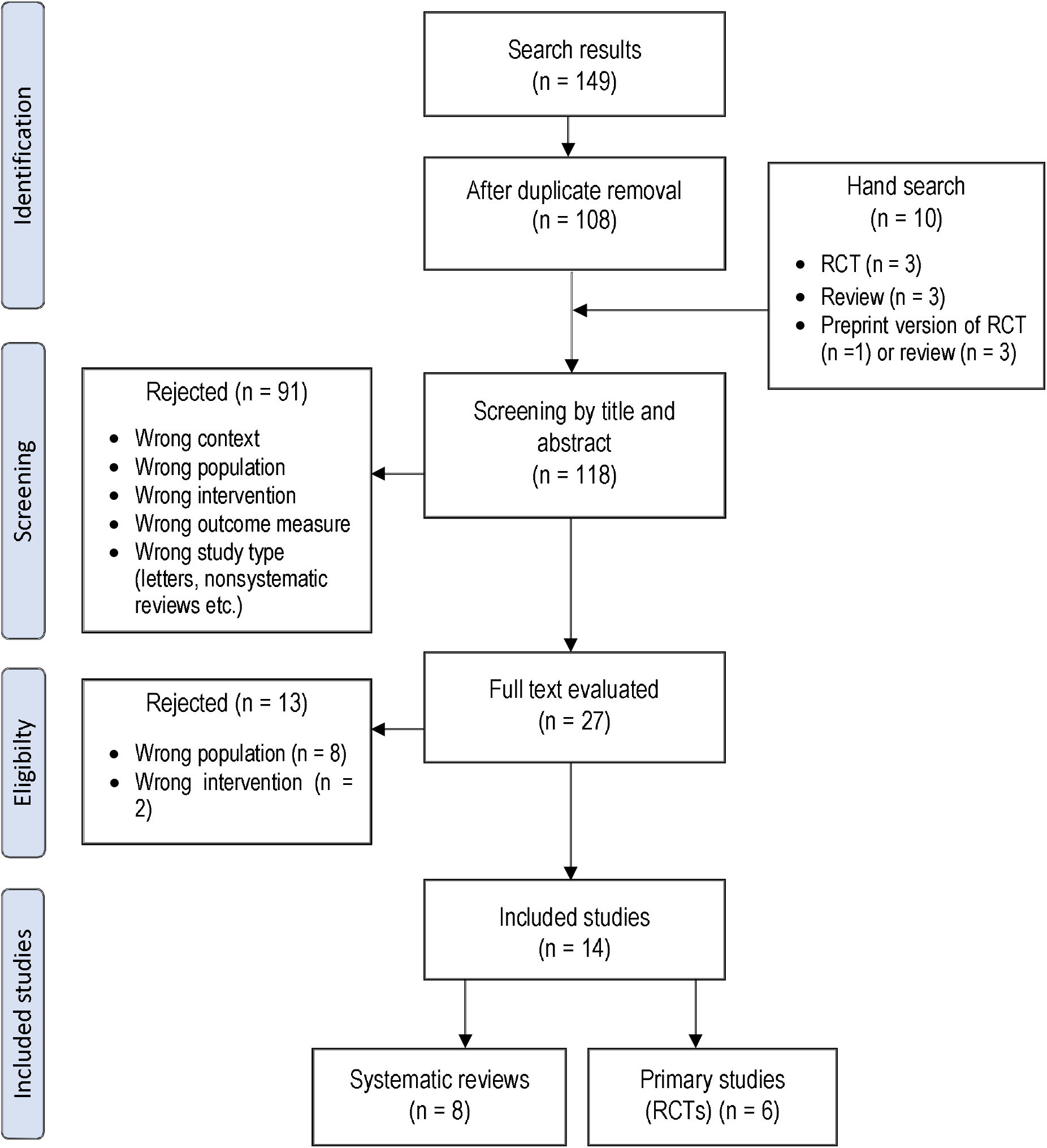
PRISMA flowchart.

In addition, until 16 May 2020, we hand searched the latest publications from the BMJ, JAMA, and the Lancet as well as the medRxiv website, which typically provides preprints of manuscripts prior to publication. We corresponded with the authors of preprints to obtain any updates to results and, where possible, a final version of the article. Background information on COVID-19 infections, methods used to diagnose and control them, and other related topics was obtained from overviews (2-4) and websites (5-7).

### Inclusion and exclusion criteria

Based on titles and abstracts, two researchers selected relevant randomized controlled trials (RCTs) and systematic reviews that corresponded to the PICO question. Articles in Danish, English, Finnish, French, German, Norwegian, Spanish, and Swedish were accepted. Observational studies, non-systematic reviews, and studies undertaken in healthcare settings or in households were excluded. Full texts of the remaining articles were read, and studies were included based on predefined inclusion and exclusion criteria based on the PICO question.

Systematic reviews were included to locate additional trials and to compare their results to our own. Simulation studies that tested masks under laboratory conditions were excluded. Cost information from included studies was to be reported, but separate searches pertaining to costs were not undertaken. Both individual and population-level results were accepted.

### Study selection

Two researchers independently assessed the risk of bias (RoB) of included publications; disagreements were resolved by a third researcher. The quality of randomized studies (n = 6) was assessed using the Cochrane Risk of Bias tool (8) with five risk-of-bias dimensions. The quality of systematic literature reviews (n = 8) was assessed by eight risk of bias questions. The quality assessment results are presented in detail in Tables 2 and 3 using the Cochrane collaboration tool (9).

**Table 2.**
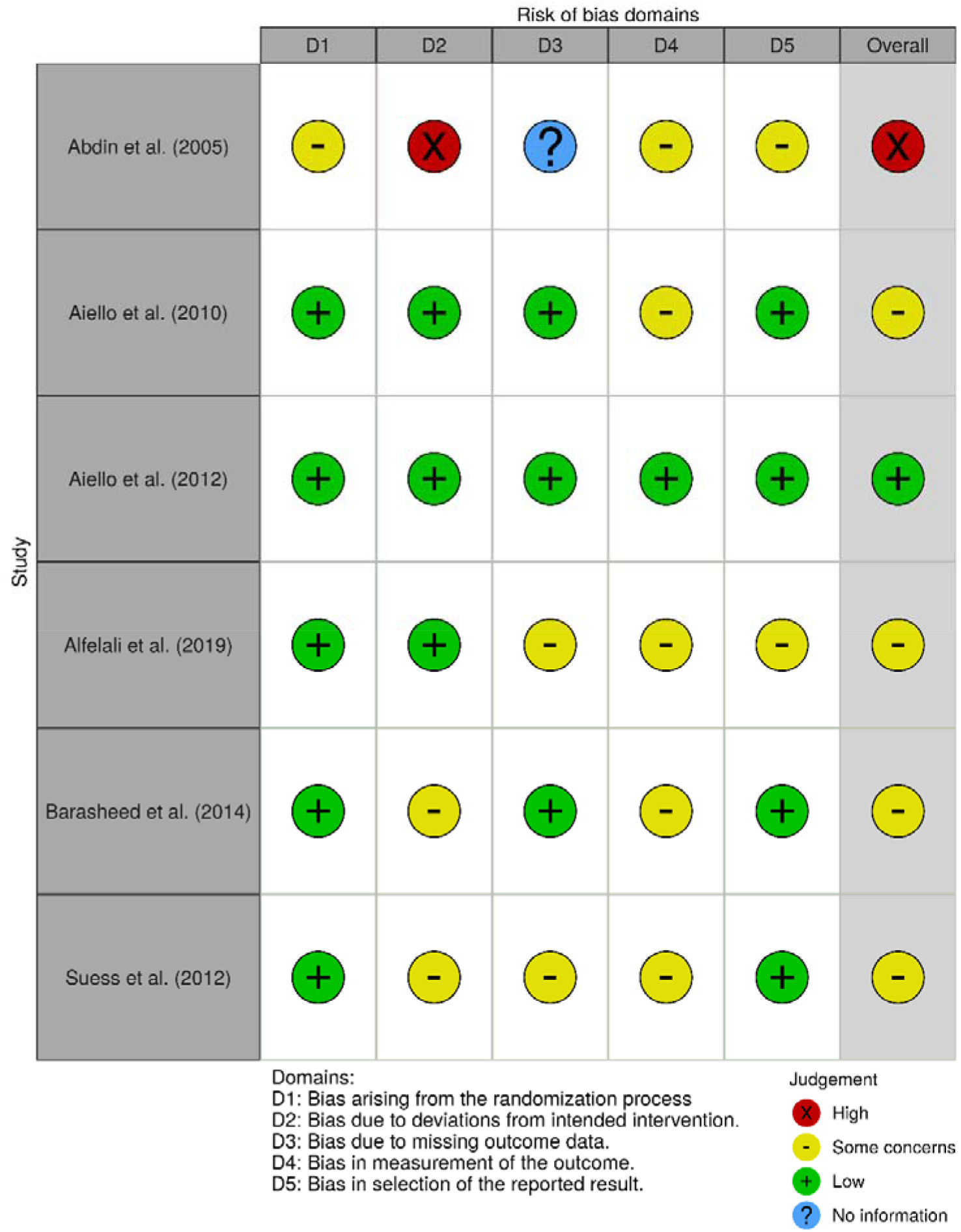
The risk of bias of included randomized trials

**Table 3.**
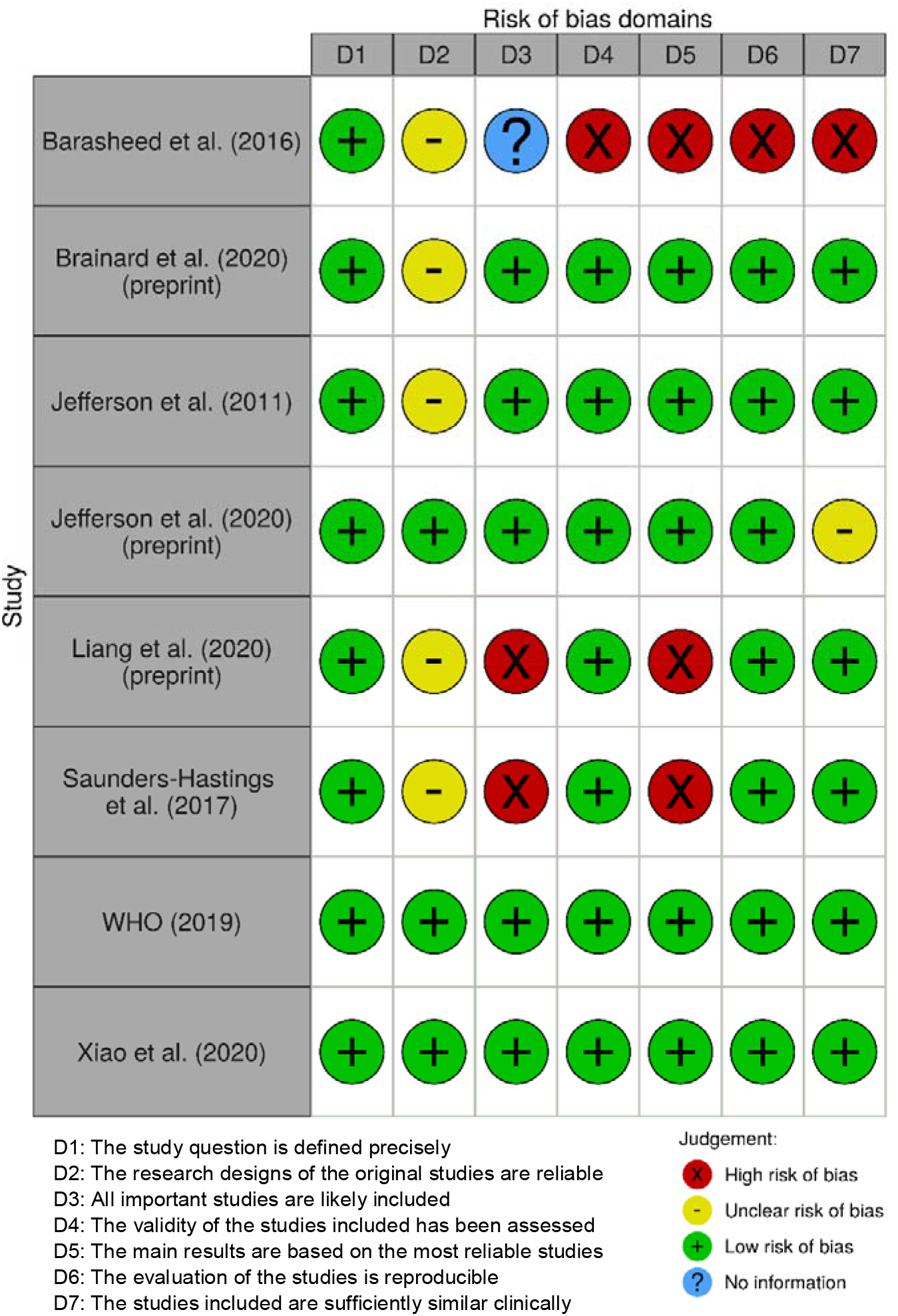
The risk of bias of systematic reviews

### Analyses of effectiveness and harms

The detailed data extracted from the original studies were tabulated separately for description of the studies (Appendix 2) and their results (Appendix 3). The results are summarized in Table 4. From systematic reviews we tabulated information on the use of face masks (Appendix 4). The tabulations were made by two of the authors, with entries by one checked by the other. The type of infection and epidemic situation were tabulated if reported.

**Table 4.**
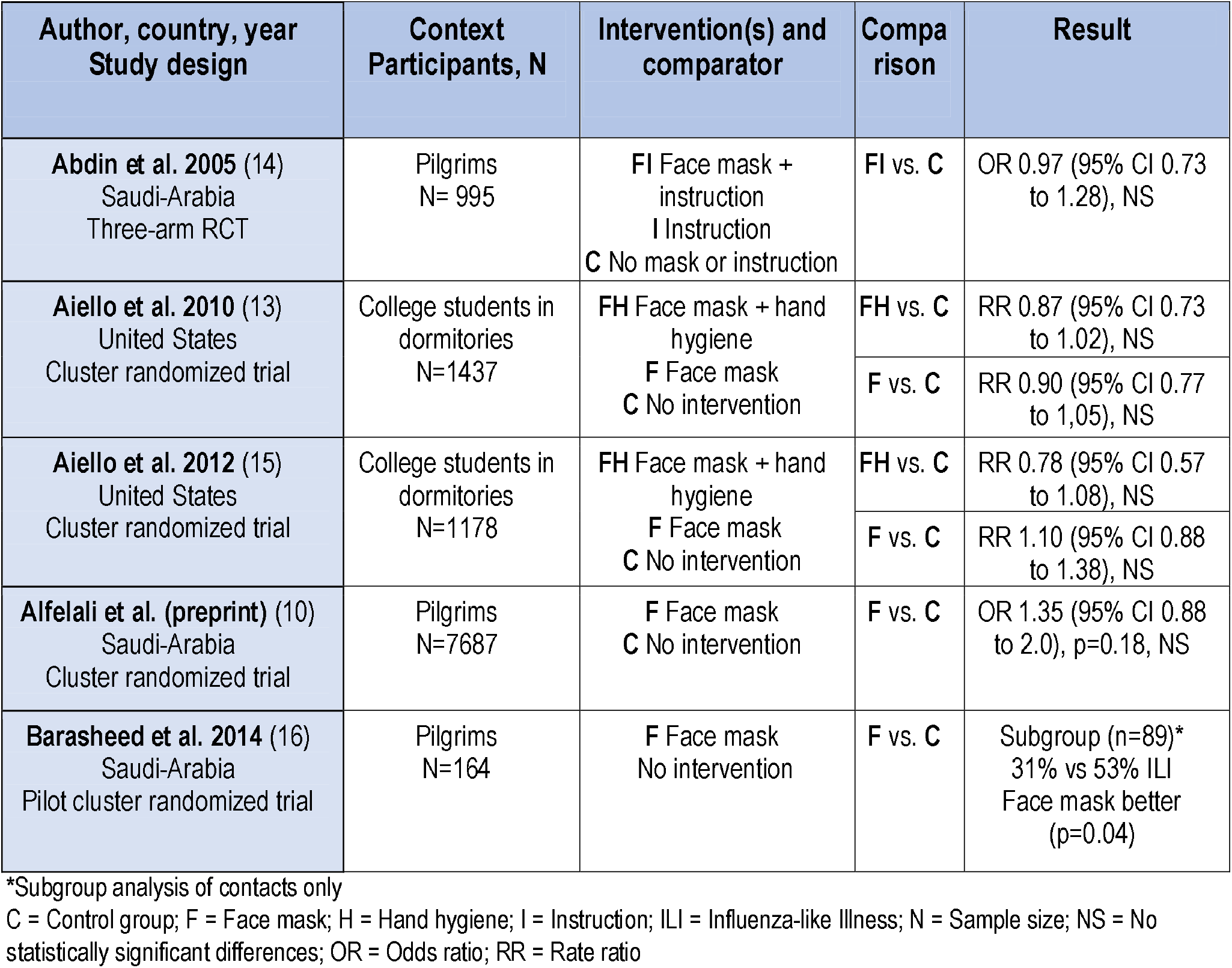
Summary of randomized trials. Detailed results can be found in Appendix 3.

Information on harms of mask use was described in one RCT (10) and one review (11). For more information, additional diary data was retrieved from an RCT by Suess et al (12); they reported mask use at home, so this study was not included in our analysis of efficacy.

Narrative summaries of each of the original studies and systematic reviews were compiled by one author and validated by another author. Our interpretation considered the applicability of the research results to a situation similar to COVID-19 in Finland, i.e., to a dangerous pandemic.

## Results

The evaluation of the effectiveness and safety of using face masks was based on six randomized studies (10,12-16). We also related our results to eight systematic reviews (4,11,17-22) that had evaluated mask effectiveness in preventing respiratory tract infections.

### Clinical effectiveness

The quality of included RCTs was variable; only one study (15) had avoided risks of bias in all five domains. The oldest study (14) was also judged to have the highest RoB. Two studies looked at university students living in dormitories in the United States and three at pilgrims traveling to Mecca. The reported dropout rate was low. Compliance with mask use was not always described in detail; mask use was quite common in control groups while not everyone in the mask groups used them. The outcome measures for main results varied: infections were verified by symptom diaries, participants’ own reports, or by antibody tests. We describe the studies below, the results are summarized in Table 4, and detailed results are in Appendices 3 and 4.

Abdin et al. (14) did not indicate the type of face masks given to participants. The main result was compliance with mask use. Infections were reported according to actual use of face masks, not randomization. There were 257 patients randomized to the mask group and 738 to the other groups. A total of 510 people used masks and 485 did not. After returning home from Mecca, 26% of participants developed an upper respiratory tract infection within a week. No difference in infections were observed between mask users and others.

Aiello et al. 2010 (13) randomized university students in clusters by college dormitories in the United States to use either a face mask (n=378), or a mask and enhanced hand hygiene (n=367), for six weeks during the 2006–2007 flu season. All participants received online training in hand hygiene. The mask group received standard surgical face masks which they were instructed to use outside home, and the hygiene group also received hand sanitizer. The comparison group (n=552) received only online education. The main outcome was the first influenza-like illness (ILI). During the six-week follow-up, the differences were not statistically significant.

Aiello et al. 2012 (15) repeated their study in 2007–2008. The mask group (n=420) received one surgical face mask for each day of the trial. In the mask and hand hygiene group (n=362), influenza-type infections were somewhat reduced compared to the control group (n=396, RR 0.78, 95% CI 0.57 to 1.08) but increased in the mask group (RR 1.10, 95% CI 0.88 to 1.38); changes were not statistically significant.

In a pilot study (16), Barasheed et al. provided surgical face masks to Australian pilgrims to Mecca (N=164). The study looked at mask use effectiveness and compliance. In the intervention group (n=75) masks were given to people with symptoms of ILI and to contacts sleeping in adjacent beds in the same tents. Controls (n=89) were not provided with facemasks, but general information on hygiene was given to them. Participants recorded their symptoms for up to five days and researchers made follow-up calls to ask about respiratory infections a week after returning home. Masks were used by 76% in mask-user tents and 12% in the control group: 13% used them while sleeping. The most common reason for not using masks was discomfort (15%). Mask use prevented infections to some extent. Of the healthy contacts who had used the mask for more than 8 hours a day, 3% became infected, compared to 9–20% close to those who used masks less or not at all.

Based on this pilot study (16), Alfelali et al. (10) carried out a cluster-randomized study in Mecca in 2013– 2015. This preprint publication is not peer-reviewed. Each mask group (n=3,864) participant received 50 surgical masks with written instructions for use. The control group (n=3,823) were allowed to use own masks if they wished. Mask use was at a relatively low level, as at least one mask per day was worn by 25% of those in the mask group (14% in the comparison group). Intention-to-treat analysis showed no reduction in the number of laboratory-certified or symptom-certified respiratory infections, or when comparing participants who wore masks on a daily basis to those who did not use them at all.

### Systematic reviews

The eight systematic reviews included some of the same randomized studies (Table 2), but the studies included varied. The quality of the reviews was also variable, with three (18-19,21) being of high risk of bias (Table 3). As the results in the reviews overlap, they cannot be combined without distortion, so each review is briefly summarized below. The methods and main results of the reviews are presented in Appendix 4.

#### Barasheed et al

(19) evaluated mask effectiveness at mass gatherings, reviewing 12 cross-sectional, 10 cohort, and two randomized studies and one case series. All studies were undertaken during pilgrimages and three targeted health care workers. The participants (N=12,710) were 11 to 89 years old and 63% were men. The utilization rate of face masks was measured in all studies, averaging 53.5 percent (range 0.02 to 92.8%). Ten of 13 included studies reporting effectiveness data used combinations of respiratory symptoms as outcome measure; viral infections were laboratory confirmed in one study; one study reported fever, and another cough as their primary outcome measure. Meta-analysis suggested a statistically significant protective effect against respiratory infections (RR = 0.89, 95% CI 0.84 to 0.94, p <0.01). According to the authors the use of face masks in mass gatherings seemed to be useful against some types of respiratory tract infections.

#### Brainard et al

(17) reviewed the effectiveness of goggles, face masks, and veils to prevent the transmission of respiratory infections in public. The preprint reported “12 cluster-randomized studies, three cohort studies, five case-control studies, and ten cross-sectional studies” while the tables show four cohort studies and one before-after design. Surgical masks were used in most of the studies. The main outcome measured the incidence of influenza-like illness (ILI) in 19 studies; four of these were carried out in schools, three with healthy people attending health care, one among the general population, one on an airplane, two among people working with animals, and seven in mass religious events (Hajj pilgrims in Mecca). Nine studies looked at infection prevention in homes.

Three RCTs showed weak evidence that mask use reduces the risk of ILI or respiratory symptoms very slightly (OR 0.94, 95% CI 0.75 to 1.19). In observational studies, masks had greater effect. In five RCTs both the infected patient and the family members wore a mask at home; the risk of infecting family members was reduced somewhat (OR 0.81, 95% CI 0.48 to 1.37), but not statistically significantly so. The protective effect was very small if mask was used only by the infected patient (OR 0.95, 95% CI 0.53 to 1.72) or only by healthy family members (OR 0.93, 95% CI 0.68 to 1.28). Authors conclude that evidence does not support routine, widespread use of face masks in the population; for high-risk individuals, mask use in short-term transient exposure may be justified.

#### The 2011 Cochrane review

(20) of non-drug interventions to reduce the transmission of respiratory viruses included 67 randomized, cohort, case-control, case-series, or before-after studies. The risk of bias was high in all five randomized and most cluster-randomized studies. The quality of observational studies varied. Only case-control studies were deemed sufficiently similar to be included in the meta-analysis.

Five trials of face masks were included, three of which showed no effect of mask use and two had some effect when combined with hand washing. Studies were not quantitatively pooled by meta-analysis due to differences in context, e.g., some studies were from hospitals. Authors recommended hand washing, masks, and isolation of infectious persons but warned that routine long-term use of some measures could be difficult to maintain outside the context of epidemics or pandemics.

A preprint version of a forthcoming **Cochrane review** (11) updates the previous one (20). Only studies looking at use of face masks as a separate measure, without support from hand hygiene and physical distancing, were included in the update. Nine studies compared the use of masks to not using them. Four of these were undertaken in homes, two in student dormitories, two in health care settings, and one during pilgrimage; none took place during a pandemic. Most studies had shortcomings in study design or reporting. Using masks did not statistically significantly reduce the incidence of influenza-like illness (RR = 0.93, 95% CI 0.83 to 1.05) or influenza cases (RR = 0.84, 95% CI 0.61 to 1.17) compared to not using a mask; no effect was seen among healthcare workers either (RR = 0.37, 95% CI 0.05 to 2.50). However, referring to their previous review, the authors recommend using a face mask in combination with other anti-infection measures.

A systematic review published as a preprint (18) by **Liang et al**. assessed whether face masks, protect against laboratory-confirmed transmission of respiratory viruses based on 13 case-control, 6 cluster-randomized and 2 cohort studies. According to the meta-analysis, mask use had a statistically significant protective effect (OR = 0.35, 95% CI = 0.24 to 0.51). Mask use protected health care workers in particular (OR = 0.20, 95% CI = 0.11 to 0.37), whereas the effect in the general population was smaller (OR = 0.53, 95% CI = 0.36 to 0.79). The protective effect appeared to be greater in Asia than in Western countries and better against SARS viruses than influenza. More than half of the studies in the review concerned health care workers. The combined results from the population studies (N=2866) is partly due to the largest study included ((23), N = 1,831), which studied bone marrow transplant patients in hospitals, i.e., not an otherwise relatively healthy general population.

#### Saunders-Hastings et al

(21) evaluated how personal protective equipment prevents the spread of pandemic influenza in the population. They found 16 studies, eight of which evaluated the effectiveness of face masks in preventing swine flu A (H1N1). In a meta-analysis of three case-control studies, the use of masks protected users from influenza, but the result was not statistically significant (OR = 0.53; 95% CI 0.16 to 1.71, p = 0.29). From the Suess study (12), the review had selected a subgroup analysis that showed a protective effect although the main result of this study was negative (OR = 0.45; 95% CI 0.2 to 1.1, p = 0.07), as the authors acknowledged. According to them, masks could be effective in future pandemics.

#### The World Health Organization

undertook a systematic review on the effectiveness of non-drug interventions in influenza epidemics to provide a basis for their guideline (4). The section on face masks worn by the population found 10 randomized studies with over 6,000 participants; six of these were in households and one in hospitals. Most studies combined mask use with measures to improve hand hygiene. Combined results showed some reduction in the risk of laboratory-confirmed infections (RR 0.78, 95% CI 0.51 to 1.20) but the results were not statistically significant (p = 0.25). The authors concluded that “there was no evidence that face masks are effective in reducing transmission of laboratory-confirmed influenza”.

#### Xiao et al. (2020)

(22) reviewed the effectiveness of non-drug interventions in preventing influenza. Their meta-analysis combined seven randomized studies with altogether 3,495 persons in face mask groups and 3052 controls. Two studies were undertaken in college dormitories, one on pilgrims, and four in households. Masks did not significantly reduce the transmission of laboratory-confirmed influenza (RR 0.78, 95% CI 0.51 to 1.20, p = 0.25), and combining adding hand hygiene did not help (RR 0.79, 95% CI 0.73 to 1.13, p = 0.39). The authors point out there is only limited evidence of the effectiveness of face masks.

### Safety of face masks

Possible harms of face masks were not systematically reported by most original studies. Alfelali et al. (10) had asked participants to record side effects on a daily basis. The most commonly reported side effect of mask use was “difficulty in breathing” (26%). Other harms were discomfort (22%); heat, sweating, bad smell, or blurred spectacles (3%); restriction of social interaction (3%) and rash (1%).

For additional information of harms, we included an RCT by Suess et al. (12) in which participants (n=172) had kept a symptom diary. This study was conducted at influenza patient’s homes in Germany during the 2009 pandemic and the 2010 influenza season. More than a third (38%) of face mask users experienced some form of harm, children (50%) more often than adults (29%, p = 0.005). The most common adverse effect was heat / humidity (children 53%, adults 35%); shortness of breath and pain were more uncommon.

Overall face masks appear to cause discomfort to users, but not actual harm. Most commonly users experienced discomfort, heat, or difficulties in breathing normally.

## Discussion

This review was commissioned and funded by the Ministry of Social Affairs and Health in Finland to inform policy decisions. We applied standard methods of systematic literature reviews and report results transparently. To find the most reliable information, the scope of our review was narrowed down to randomized trials examining the effects of out-of-home use of face masks on the transmission of droplet-mediated respiratory infections.

The comprehensiveness of our literature search was ascertained by reviewing the reference lists from systematic reviews on the topic and from recent recommendations by the WHO (4) and the European Center for Disease Prevention and Control (7). In addition, we followed new publications and online discussion in key scientific journals, as we had exceptionally agreed to include non-peer-reviewed publications (preprints), as COVID-19 is being actively studied.

The large study by Alfelali et al (10) is not peer-reviewed. Three non-peer-reviewed reviews were included, all of which were of reasonably low risk of bias. In one of these (18), results with better efficacy were selected from subgroups of an included study instead of presenting the main outcome of the study.

The studies had not always monitored the effect of masks on transmission of infections, but looked at the symptoms (cough, fever, etc.) and thus could not identify the disease of interest from other infections. Some studies looked at the acceptability of using masks and reported infections poorly. The included studies did not provide sufficient detail to gauge possibly increased disease transmission via inexperienced mask use, e.g., adjustment of face coverings when wearing them, inappropriate disposal or cleaning, or reduced adherence to physical distancing and hand-hygiene guidance.

Our research question covered several different types of masks, user groups, and operating environments outside health care. We did not combine the results of the randomized studies in meta-analysis as the studies were few and their target populations differed.

### Results of randomized trials

Four of the randomized studies (10,13,15-16) reported the effect of masks on transmission of respiratory infections and from the fifth study (14) it was possible to calculate the effect using the data provided (Table 4, Appendix 3). The use of masks prevented infections in a subgroup of one pilot study (16). Due to the heterogeneity of the data, it was not appropriate to perform a meta-analysis. The quality of RCTs varied and due to the nature of the intervention, the studies were not blinded. Possible blinding of the data analysis was not mentioned.

Most randomized studies used commercial surgical masks. Standards for face masks are based on their effectiveness in filtering particles in laboratory tests. The mask should apparently be changed often enough, and cloth face coverings cleaned between uses; these measures are rarely reported in original studies. There was no information in the studies on possible transmission of infections from used masks.

None of the original trials looked at the use of face masks in situations resembling urban environments in Northern Europe. Three were made in Mecca during the pilgrimage, where participants spent several days in crowded conditions, and two in the United States, in student dormitories on campus during a period of seasonal influenza. The applicability of the studies to urban populations is difficult to assess; the crowding during pilgrimage seems quite different infectious environment than a European shopping center or public transportation.

Adherence to using a face mask could be assessed by recording what proportion of the population wear masks, how much of the time they wear them, and how often masks are changed. Some studies measured use by objective observation, others with diaries or questionnaires, and some studies did not report use or adherence at all. The rate of wearing masks varied widely between studies, and it was quite common for control participants to wear masks.

The effect of masks on infection prevention was measured in a variety of ways in the controlled trials included in our review. Follow-up periods ranged from a few days to several weeks. Virus or antibody assays had been used infrequently and mostly in symptomatic participants only. Several studies used symptom combinations to identify an acute respiratory infection or influenza-like infection (ILI). Some identified illness based on a single symptom (e.g., fever or cough).

Immunity acquired during previous infections may also have reduced the number of infections, whereas there is currently thought to be little immunity to SARS-CoV-2 in populations. The modes of transmission of this virus and the barrier function of face masks against it are not well known yet.

Based on randomized studies, the effect of face masks on the transmission of upper respiratory tract infections outside the home appears small or nonexistent. The cost or cost-effectiveness of using face masks was not reported in any of the original studies included. The provision of free masks increased their use in one study (14).

### Results of reviews

The results of this review are in line with earlier reviews: relevant research is scant, and the effect of face masks in preventing transmission of respiratory infections in the community is not backed up by strong evidence. Masks are rarely used as the only public health measure in epidemics or pandemics, so their independent effect on transmission is especially difficult to evaluate.

The quality of the methods in the systematic reviews varied and in some reviews the risk of bias was significant. The most common source of bias was acceptance of weak study designs. The authors of the reviews themselves often used arguments other than those arising from the results of the review, such as the ease of non-drug interventions. One review (18) analyzed the risk of publication bias using a funnel plot and bias appeared to be slight. In publications analyzed in the reviews, mask type was not always defined. Hand washing and education were used as additional measures in many studies.

The reviews (4,11,17-22) show that the effect of the use of face masks on transmission of infections has not been reliably demonstrated. Five of the eight systematic reviews showed no evidence of face masks being helpful in controlling the source of respiratory infection or preventing exposure in healthy individuals. Three reviews were cautiously positive; one (19) concerned pilgrims in Mecca and another (18) included bone marrow transplant patients in hospital. The positive result of one review (21) was based on favorable partial results extracted from the data. Three of the reviews (11,17-18) were in preprint form, i.e. had not yet been peer-reviewed.

Each of the eight reviews included at least one randomized study located by our review, but none had found them all. In most cases, the reviews had accepted other study designs with higher risk of bias and included both health care and other types of environments. Meta-analyses often combined very different studies.

Some authors’ conclusions seem to be based on thinking that protection by non-drug measures against respiratory infections is “mechanically plausible”, despite their own results showing little or no effect. Some reviews commented on the relative affordability of commercial face coverings, but the costs or cost-effectiveness of using masks were not reported. Brainard et al. (17) suggested that the balance of benefits and costs of using masks to prevent disease should be estimated.

After our review was published in Finland in May 2020 (24), Chu et al. (25) published a systematic review and meta-analysis investigating the optimum distance for avoiding transmission of SARS and Middle East respiratory syndrome viruses and to assess the use of face masks and eye protection to prevent transmission of these viruses. They included study designs of all types and found three original retrospective studies on the use of face masks outside health care in China or Vietnam. Two of these (26-27) looked at close household and community contacts of SARS patients, and one (28) was a case-control study of SARS patients who had no reported contact with other SARS patients. Wearing masks had a protective effect in all three studies. These results indicate that the use of masks in populations that are used to wearing them may add to other public health measures during a SARS virus epidemic.

A new randomized study with 3030 participants from Denmark was published in November while our study was under review (29). Infection with SARS-CoV-2 occurred in 1.8% of participants in the mask group and in 2.1% of controls, amounting to a difference of −0.3 percentage point (95% CI, −1.2 to 0.4; P = 0.38). This trial from European context is in line with earlier results.

## Conclusions

There are very few randomized studies on the effect of face masks on the transmission of respiratory infections in the general population. According to the trials and systematic reviews found by our review, the effect of face masks used outside the home on transmission of droplet-mediated respiratory infections in the population is minimal or non-existent. It is difficult to distinguish the potential effect of face coverings in trials from the effects of other protective measures.

There are few reliable studies on the subject and their target populations and research environments clearly differ from a general urban population. No randomized studies have been performed on the efficacy of non-surgical (self-made or commercial) face masks in the general population. We did not find information of the cost or cost-effectiveness of using face masks; the time available did not allow for a targeted literature search on costs.

It is difficult to apply our results to the COVID-19 epidemic, as the public use of face masks would take place in quite different conditions from those studied, and the virulence of SARS-CoV-2 differs from infections studied. In order to assess real-world effectiveness, information on the level of adherence to the use of masks in each country would also be needed.

The absence of evidence of the effects of an intervention does not necessarily mean that the intervention is not effective. However, the effectiveness of face masks has been studied in thousands of people, so a clearly protective effect could be expected to have emerged.

## Data Availability

All data relevant to the study are included in the article or uploaded as supplementary information

